# Assessing anterior shoulder instability: Reliability of the humeral head centralization test

**DOI:** 10.64898/2026.11.09.26367343

**Authors:** Marianne L. van Gastel, Karin M.C. Hekman, Bouke J.H. Boden, Gwendolyne G.M. Scholten-Peeters, Lea M. Dijksman, Michel P.J. van den Bekerom

**Affiliations:** Department of Physical Therapy, Ziekenhuis Amstelland, The Netherlands; Department of Orthopedic Surgery, Medical Centre Jan van Goyen, The Netherlands; Department of Radiology, Medical Centre Jan van Goyen, The Netherlands; Department of Human Movement Sciences, Vrije Universiteit Amsterdam, Amsterdam, The Netherlands; Department of research and epidemiology, St. Antonius Hospital, Nieuwegein, The Netherlands; Shoulder and Elbow Unit, Department of Orthopedic Surgery, OLVG, Amsterdam, The Netherlands

**Keywords:** Rehabilitation, Shoulder, Joint Dislocation, Diagnostic Imaging, Reproducibility

## Abstract

**Background:** Motor control impairment of the rotator cuff muscles is reported to compromise the stability of the glenohumeral joint. However, an easily applicable, reliable parameter for quantifying anterior translation of the humeral head during active arm movements is lacking. Therefore, a novel (humeral head centralization) test was developed and evaluated.

**Study design:** Case-control study with reliability analysis (Level III).

**Methods:** This research project examined the shoulders of 25 patients with anterior shoulder instability and 25 asymptomatic controls. A physical therapist assessed humeral head translation in both groups during shoulder external rotation and elbow extension using ultrasound imaging. For test-retest and inter-rater reliability, the ultrasound assessment was repeated by the physical therapist and a radiologist after one week. Patients, the physical therapist, and the radiologist were blinded to the test results. In the evaluation of the difference in translation between the patients and controls, the Wilcoxon rank sum test was employed. The test-retest and inter-rater reliability were estimated using Intraclass correlation coefficients (ICC_2,1_).

**Results:** In patients, in active elbow extension, a significantly greater anterior translation of the humeral head was observed compared to participants in the asymptomatic group (p < .001). The test-retest reliability of the physical therapist demonstrated poor agreement for the external rotation movement (ICC_2,1_ 0.32 (95% CI -0.08-0.63)) and moderate agreement for the extension movement of the elbow (ICC_2,1_ 0.66 (95% CI 0.36-0.84)).

The inter-rater reliability of the physical therapist and radiologist was moderate for both the external rotation (ICC_2,1_ 0.51 (95% CI 0.16-0.75)) and elbow extension movements (ICC_2,1_ 0.62 (95% CI 0.30-0.82)).

**Conclusion:** Although translation of the humeral head during active elbow extension movement was increased in patients compared to controls, this study found difficulties in reliably measuring anterior glenohumeral translation. Therefore, this novel test, in its current form, cannot be recommended for use in clinical practice or research.

## Introduction

The incidence of anterior shoulder dislocations is between 21 and 27 per 100.000 person-years. (1),(2),(3)

A traumatic anterior dislocation of the glenohumeral joint may result in lesions to the glenoid bone or the humeral head as well as damage to the capsulo-labral complex. Lesion to the capsulo-labral complex may again result in a diminishment of negative intra-articular pressure, which may disturb joint mechanics and motor coordination.(4) Additionally, pain, apprehension, capsulo-ligamentous pathoanatomy, and impaired neuromuscular control may alter kinematics during functional tasks.(3) Following the initial dislocation, recurrent instability has been reported in 19-88% of patients, with patient characteristics, bony lesions, and motor control deficiency identified as factors related to recurrence.(5),(6),(7), A reduction in sensorimotor function of the capsuloligamentous structures, resulting from injury and abnormal muscle activation patterns, has been associated with impaired dynamic stabilization provided by the rotator cuff.(8),(9)

In managing and restoring anterior glenohumeral stability, shared decision-making relies on patient characteristics such as age, sex, sport type and intensity, and bone loss.(10) However, information regarding the dynamic stability of the glenohumeral joint is lacking.

The inability to actively control humeral head centralization during movement may serve as an indicator of a changed direction of compression force, aberrant movements and/or motor control deficiency following dislocation. Previous ultrasound and stress radiography studies showed that patients with anterior shoulder instability exhibit greater anterior translation under anterior stress compared to asymptomatic shoulders.(11),(12),(13)

Ultrasound is a valid and reproducible method for assessing shoulder laxity.(14) The use of ultrasound to assess humeral head translation during supported arm movements has the potential to provide a non-invasive method that is both clinically practical and learnable.

To the best of our knowledge, ultrasound has not previously been used in the evaluation of the humeral head position during active movements in the anterior unstable shoulder.

A case-control study with inter-rater and test-retest reliability analysis was conducted. The Strengthening the Reporting of Observational studies in Epidemiology (STROBE) guidelines and the Guidelines for Reporting Reliability and Agreement Studies (GRRAS) were used.(15),(16) The Medical Research Ethics Committee United (MEC-U) approved the study (NL69882.100.19.). All participants signed informed consent prior to inclusion in the study.

### Participants

Between the 31^st^ of May 2021 and 13^st^ January 2025, 25 consecutive patients were included from the orthopedic department. Additionally, 25 asymptomatic matched participants were included. Eligible patients were at least 18 years old, had recurrent instability (at least two involuntary re-dislocations or subluxations) following anterior shoulder dislocation, and opted against immediate surgical intervention. Patients needed to be able to read and write in Dutch or English to complete the questionnaires and to sign informed consent. Patients were excluded in cases of generalized hyperlaxity (Beighton score >6 points), atraumatic, posterior, or multidirectional instability, or those with neurological deficits. Patients who sustained a bony lesion (shown on conventional radiographs) due to dislocation were also excluded, as were patients who had undergone stabilizing surgery of the affected shoulder. The baseline characteristics, patient-reported outcome measures, and results of physical testing are shown in **Table 1**.

**Table 1.**
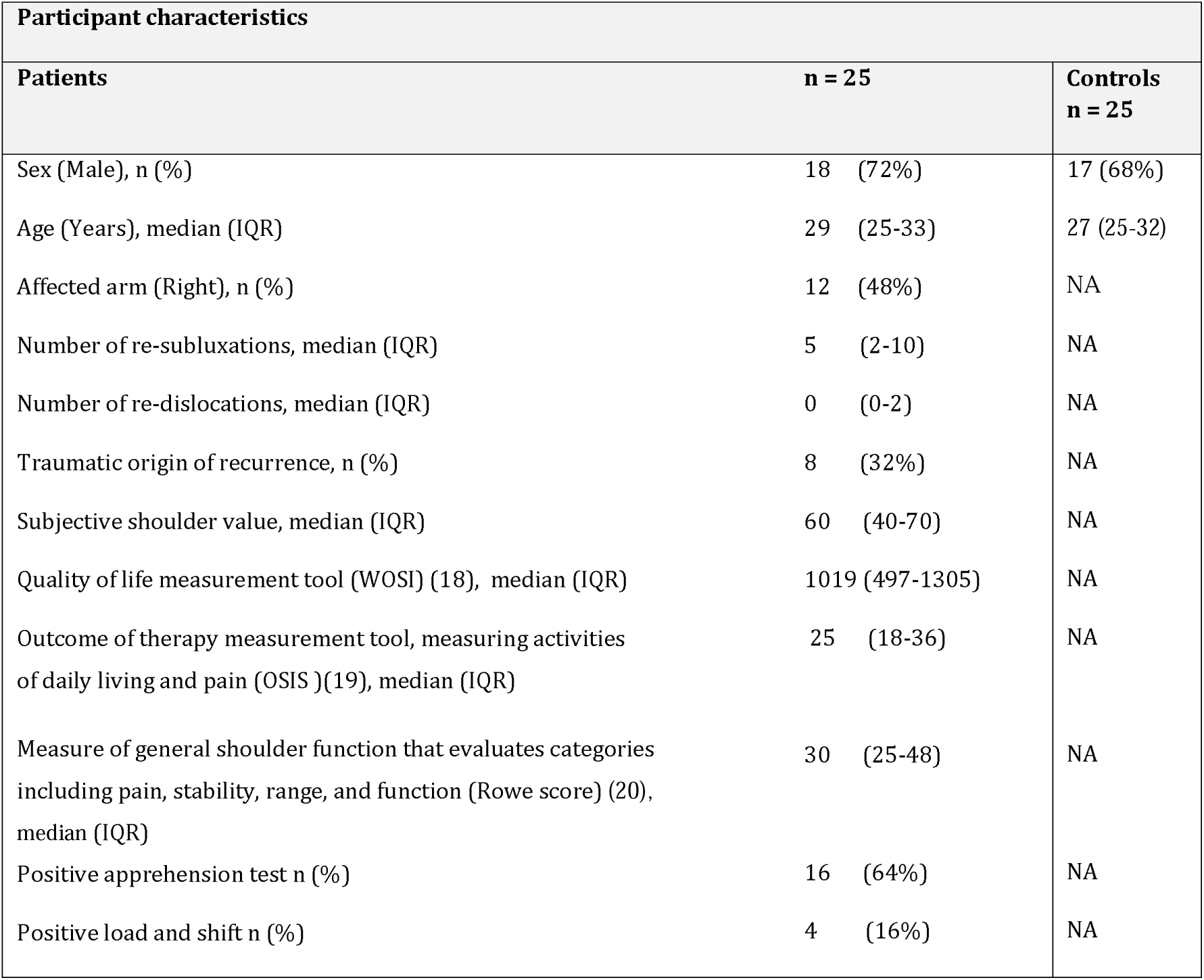
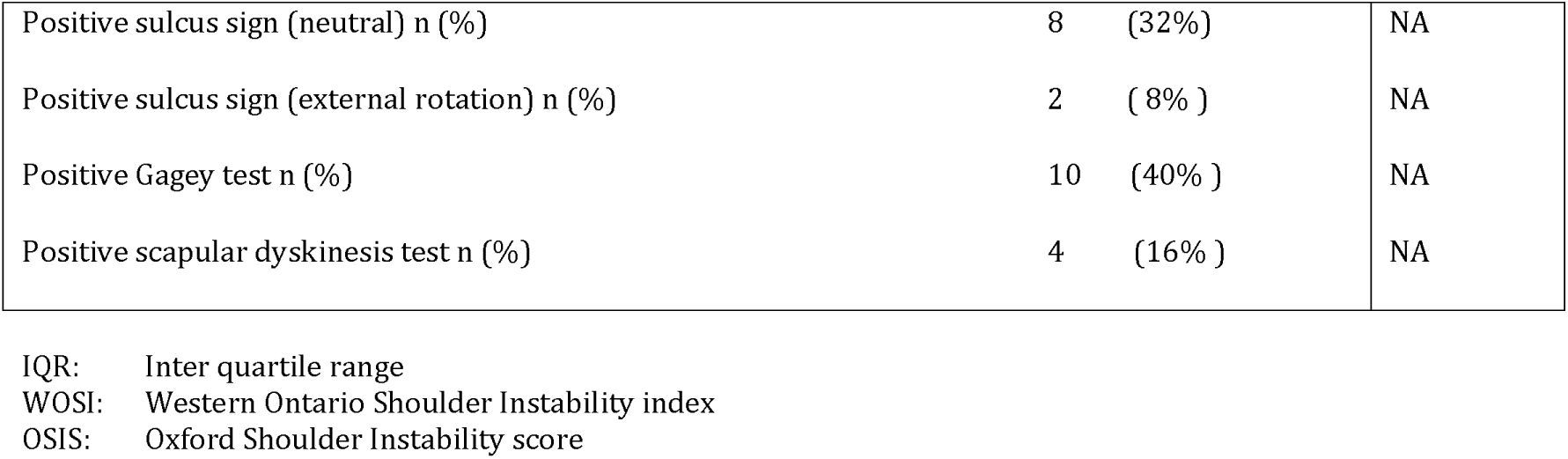
Characteristics of participants n = 50.

| Participant characteristics |  |  |
| Patients | n = 25 | Controls<br>n = 25 |
|---|---|---|
| Sex (Male), n (%) | 18 (72%) | 17 (68%) |
| Age (Years), median (IQR) | 29 (25-33) | 27 (25-32) |
| Affected arm (Right), n (%) | 12 (48%) | NA |
| Number of re-subluxations, median (IQR) | 5 (2-10) | NA |
| Number of re-dislocations, median (IQR) | 0 (0-2) | NA |
| Traumatic origin of recurrence, n (%) | 8 (32%) | NA |
| Subjective shoulder value, median (IQR) | 60 (40-70) | NA |
| Quality of life measurement tool (WOSI) (18), median (IQR) | 1019 (497-1305) | NA |
| Outcome of therapy measurement tool, measuring activities of daily living and pain (OSIS )(19), median (IQR) | 25 (18-36) | NA |
| Measure of general shoulder function that evaluates categories including pain, stability, range, and function (Rowe score) (20), median (IQR) | 30 (25-48) | NA |
| Positive apprehension test n (%) | 16 (64%) | NA |
| Positive load and shift n (%) | 4 (16%) | NA |
| Positive sulcus sign (neutral) n (%) | 8 (32%) | NA |
| Positive sulcus sign (external rotation) n (%) | 2 ( 8% ) | NA |
| Positive Gagey test n (%) | 10 (40% ) | NA |
| Positive scapular dyskinesis test n (%) | 4 (16% ) | NA |

IQR: Inter quartile range
WOSI: Western Ontario Shoulder Instability index
OSIS: Oxford Shoulder Instability score

### Procedure

All patients were asked to complete a set of patient-reported outcome measures, including the Western Ontario Shoulder Instability index (WOSI), the Oxford Shoulder Instability score (OSIS), and the Simple Shoulder Value. Secondly, a physical examination of the affected shoulder was performed. Finally, an ultrasound assessment (Humeral Head Centralization test) of the translation of the humeral head during movement was performed. The physical examination and ultrasound assessment were performed by the physical therapist (KH). After one week, during which the patient did not receive any treatment, the ultrasound assessment was repeated by the physical therapist (KH) and, within ten minutes, again by a radiologist (BB).

The control group individuals underwent a single ultrasound examination of their dominant shoulder performed by the same physical therapist (KH).(**Figure 1**)

**Figure 1.** Flowchart of assessment

### Raters

A physical therapist (KH) with 20 years of primary/secondary care experience and at least seven years of expertise in musculoskeletal ultrasound performed the first and second shoulder ultrasound examinations. She holds a certificate in “Upper Extremity MSU Skills” and an MSc in Manual Therapy, and conducts approximately 200 shoulder ultrasound scans annually in primary care. A radiologist (BB) with over 12 years of experience as a musculoskeletal radiologist performed the second ultrasound examination. His background includes a one-year fellowship focused on musculoskeletal ultrasound and 16 years of experience in shoulder ultrasound assessment. He performs more than 1,000 shoulder ultrasound examinations annually.

### Radiological examination

A Philips Affiniti 70G (Hardware Version 6.0.0) ultrasound scanner was used to assess humeral translation. The HHC test was performed with the patient or control group participant lying prone. The shoulder was positioned at 90° of abduction, supported by towels in a stable position to prevent the upper arm from moving during the test movements. The ultrasound probe was positioned right beneath the posterior edge of the acromion in a transverse position with the posterior glenoid rim and the humeral head visible. This starting position was measured and saved. (**Figure 2**)

**Figure 2.** Illustration of the starting position of the test. Prone patient positioning with supported upper arm: shoulder at 90° abduction and in neutral rotation. A depicts the posterior edge of the humeral head in the starting position, B the posterior glenoid rim

A perpendicular horizontal line was placed at the posterior edge of the humeral head (A) and the posterior glenoid rim (B). The distance between the horizontal lines, A and B, was measured as a positive (+) number. When the posterior humeral head (C) was situated anterior to the glenoid rim line (B), the measured distance was given a negative number (-) **Figure 3**).

**Figure 3.** Illustration of the hypothesized translation of the humeral head in mm. It shows the hypothesized direction of humeral head translation during maximal active external rotation and/or elbow extension in the presence of motor control impairment. A depicts the posterior edge of the humeral head in the starting position, B the posterior glenoid rim, C the posterior edge of the humeral head during movement.

Three ultrasound measurements were conducted, all initiated from the same starting position of 90 degrees of abduction and 90 degrees of elbow flexion in the prone position, with the thumb pointing towards the plinth. All participants performed an identical series of movements.

1. Starting position: Shoulder supported in 90 degrees of abduction and 90 degrees of elbow flexion in the prone position, with the thumb pointing towards the plinth (**Figure 4**).

**Figure 4.** Starting position

2. Test movement 1: The patient was instructed to perform a full external rotation of the shoulder, with the thumb oriented toward the plinth. At the end of this external rotation, the image was recorded, and the humeral position was measured (**Figure 5**).

**Figure 5.** Test movement 1

3. Test movement 2: The patient was instructed to perform a full elbow extension (the thumb oriented toward the ground). The image was recorded, and the humeral position was measured at full extension (**Figure 6**).

**Figure 6.** Test movement 2

The starting position and test movements were selected based on clinical experience, injury mechanisms in traumatic anterior shoulder instability, and the practicality of the measurement. In the patient group, the examination was focused on the affected shoulder, whereas in the control group, the examination was conducted on the dominant side. For familiarization with the two test movements, the patient or control participant was instructed to perform four repetitions of the movement prior to recording. The result of the fifth movement was measured and recorded.

The raters were comprehensively trained before and continuously during the inclusion period to pursue agreement in the execution of the Humeral Head Centralization test

### Blinding

The patients were blinded to the repeated ultrasound measurements. The physical therapist and radiologist were blinded to the results of the previous measurements. The measurements of both raters were conducted independently.

### Sample size calculation

A priori sample size analysis was performed using the PS Power and Sample Size Calculator (version a57e8c3, Universität Vanderbilt University, Tennessee). To detect a true mean difference of 3.00 mm between the experimental and control group, with a standard deviation of 3.50 mm(12), a sample of 23 participants per group was required to achieve 80% power at a two-sided significance level of 0.05. Taking a dropout rate of 10 % into account 25 patients per group were needed.

### Statistical analysis

Descriptive statistics were used to summarize demographic data. All continuous data were described using mean and standard deviation or median and interquartile range (IQR), when the data were not normally distributed. Categorical variables were described as frequencies and percentages.

The difference in the HHC test results between the patients and the controls was assessed using a Wilcoxon rank sum test (non-parametric). The test-retest reliability and the inter-rater reliability of the HHC test was calculated using the ICC_2,1_ (single measures, two-way random effect, absolute agreement). The ICC_2.1_ values were interpreted according to Koo and Li, in which an intraclass correlation coefficient (ICC) < 0.50 was considered poor reliability, 0.50 to 0.75 moderate reliability, 0.76 to 0.90 good reliability, and >.90 excellent reliability.(17) All ICCs were reported with 95% confidence intervals (CI). Moreover, the standard error of measurement (SEM) was calculated using the formula *SEM = SD ×* √*(1–ICC)*.

## RESULTS

A total of 25 patients with shoulder instability and 25 asymptomatic controls were included. No patients dropped out. The median (IQR) age of the patients was 29 years (25–33), and 18 were male (72%). In comparison, the median (IQR) age of the control group was 27 years (25–32). Characteristics of the patients and controls are summarized in **Table 1**. The median (IQR) Humeral Head Centralization test in external rotation of the patients was 4.9 mm (1.1-7.6) and of the controls 7.6 mm (6.2-10.6). The median (IQR) Humeral Head Centralization test in extension of the elbow of the patients was -1.5 mm (-4.6-.6) and of the controls 2.9 mm (0.5-6.4).

### Humeral head centralization during active movement

In the patient group, the median initial position of the humeral head was measured at 6.6 mm. During active external rotation of the arm, the humeral head translated 1.7 mm anteriorly, resulting in a position 4.9 mm posterior to the glenoid rim. Furthermore, during active elbow extension, the humeral head translated from 6.6 mm posterior to the glenoid rim to a position of -1.5 mm anterior of the glenoid rim, indicating an anterior translation of 8.1 mm.

In the control group, the median initial position of the humeral head was 6.4 mm. During active external rotation of the arm, the humeral head translated 1.2 mm posteriorly, resulting in a position 7.6 mm posterior to the glenoid rim. Furthermore, during active elbow extension, the humeral head translated from 6.6 mm posterior to the glenoid rim to a position of 2.9 mm, indicating an anterior translation of 3.7 mm (**Table 2**).

**Table 2.** Humeral Head Centralization test results (in mm)

| **Examination by the physical therapist** | **1<sup>st</sup> evaluation median(IQR)** | **Single evaluation median (IQR)** | **P-value** |
|---|---|---|---|
|  | **Patients** | **Control group** |  |
| HHC test (Starting position) | 6.6 (5.3-8.2) | 6.4 mm (IQR 3.3-8.4) | .295 |
| HHC test (Active external rotation) | 4.9 (1.1-7.6) | 7.6 mm (IQR 6.2-10.6) | .006 |
| HHC test (Active elbow extension) | -1.5 (-4.6-0.6) | 2.9 mm (IQR 0.5-6.4) | < .001 |

### Differences in humeral head centralization between patients and controls

In the patient group, the median humeral position in the starting position was 6.6 mm, compared to 6.4 mm in the control group (p=.295). During active external rotation, the median position of the humeral head of the patients was 4.9 mm, compared to 7.6 mm in the control group (p=.006). During active elbow extension, the median position of the humeral head of the patients was -1.5 mm, compared to 2.9 mm of the control group (p <.001). This difference of 4.4 mm was larger than the Standard Error of Measurement (SEM) of 2.1mm.

### Test-retest reliability

The results of the second and third assessments, performed with 10 minutes in between, by the physical therapist and radiologist are summarized in **Table 3**.

**Table 3.** Humeral Head Centralization test results (in mm)

|  | **2<sup>nd</sup> and 3<sup>rd</sup> evaluation, Median (IQR)** |  |
|  | **Radiologist** | **Physical therapist** |
|---|---|---|
| Starting position | 3.5 (2.3-4.6) | 6.2 (4.4-7.8) |
| External rotation | 5.2 (1.4-6.1) | 5.5 (3.7-6.9) |
| Elbow extension | -1.5 (-4.3-.0) | -2.8 (-4.6-1.3) |

Test-retest reliability of the physical therapist’s evaluations in the starting position showed moderate agreement. In contrast, the assessment of active external rotation demonstrated poor reliability, while elbow extension showed moderate reliability (**Table 3**).

The inter-rater reliability of the results of the physical therapist and the radiologist showed poor agreement in the starting position. The reliability of the active external rotation as well as the active elbow extension, was moderate (**Table 3**).

**Table 3.**
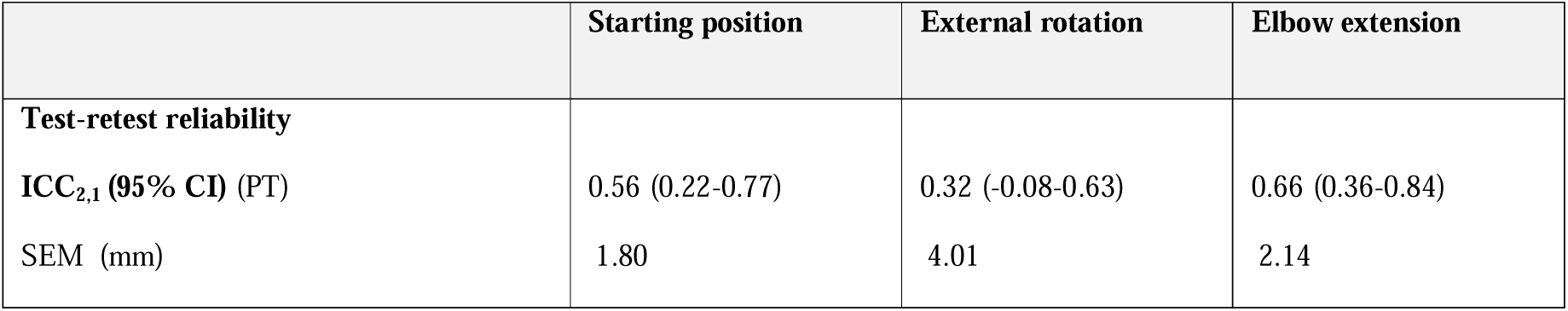
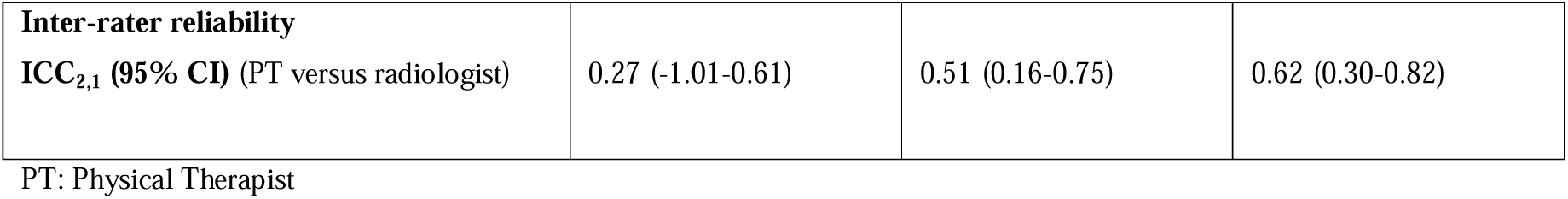
Test-retest and Inter-rater reliability shown in the Intraclass coefficient with 95% confidence interval.

|  | **Starting position** | **External rotation** | **Elbow extension** |
|---|---|---|---|
| **Test-retest reliability** |  |  |  |
| **ICC<sub>2,1</sub> (95% CI)** (PT) | 0.56 (0.22-0.77) | 0.32 (-0.08-0.63) | 0.66 (0.36-0.84) |
| SEM (mm) | 1.80 | 4.01 | 2.14 |
| **Inter-rater reliability** |  |  |  |
| **ICC<sub>2,1</sub> (95% CI)** (PT versus radiologist) | 0.27 (-1.01-0.61) | 0.51 (0.16-0.75) | 0.62 (0.30-0.82) |

PT: Physical Therapist

## DISCUSSION

This study revealed a significant difference in humeral translation between the patient group and the control group during active elbow extension, whereas no significant difference was detected during shoulder external rotation. Furthermore, we found poor to moderate reliability in terms of test-retest and inter-rater assessments.

Previous ultrasound and stress radiology research, measuring the anterior translation in the shoulder joint, also identified a difference between patients and controls in humeral translation.(12),(13) However, due to the different setup, direct comparisons of the results cannot be made. The ultrasound study, for instance, used a setup with the arm positioned in internal rotation and secured to the body, with the transducer placed anteriorly on the shoulder.(12) This study aimed to develop a reliable ultrasound test with a simple setup to enhance its feasibility for routine clinical application. However, this simple setup appeared to be associated with a poor to moderate test-retest reliability and inter-rater reliability. Again, comparing these results to recent literature is difficult due to significant differences in study design, setup, and choice of participants.(11) In a study where 39 patients were examined under anesthesia before stabilizing surgery, a good test-retest reliability and a moderate inter-rater reliability were found.(11) In another ultrasound study on humeral translation involving 12 asymptomatic individuals, the arm was fixated with straps in neutral abduction/internal rotation and in 90° abduction/external rotation using an anterior view. The study reported good to excellent test-retest reliability and moderate intra-rater reliability.(21) Another study evaluating shoulder laxity among asymptomatic individuals found a moderate test-retest reliability and an excellent inter-rater reliability.(14) The test-retest reliability for active external rotation in the present study was found to be poor. These findings suggest that testing of active external rotation in a supported arm in 90 degrees of abduction should be reconsidered and not recommended. A possible reason for this low reliability might be that the visibility of the bony landmarks, which are used in the HHC test, change due to the external rotation of the humeral head.

This test might be important given the current lack of practical, deployable measures of sensorimotor control, which are relevant in the rehabilitation of patients with shoulder instability.(9),(22) However, the study also highlights the difficulties of measuring the anterior translation of the glenohumeral joint in active motion. The results of the HHC test are influenced by multiple variables. Achieving standardization in both patient positioning and probe placement proved challenging when prioritizing an easily applicable setup. For example, a standardized support for the upper arm that did not restrict its movement was not identified. Additionally, choosing to examine the dominant side of the control group may have led to a greater disparity between groups, as the dominant side could demonstrate superior motor control and consequently less anterior translation compared to the non-dominant side. Another limitation was the extended period of 39 months of inclusion, which may have affected the consistency in performing the new ultrasound examination.

In addition to the limitations, this study also has strengths. First, the use of the posterior approach has been advocated due to its advantage of the scapular spine as a bony landmark, which can be employed to secure a reproducible transducer position. Also, it has been reported to reach higher inter-rater reliability than the anterior approach.(11),(21) Secondly, for a radiologist or a trained physical therapist, this test is straightforward to implement in routine practice for diagnostic and evaluative purposes. The test may also serve a role as a biofeedback mechanism to assist patients in actively centralizing the humeral head within the glenoid cavity. In this process of active feedback, where conscious activation of the rotator cuff is guided by the physical therapist, the reliability of the test should also be estimated.

Future research should focus on further developing the application of both ultrasound and CT analysis. When employing ultrasound, it is important to prioritize enhanced standardization without compromising its applicability. Standardization, comprehensive and repeated training for both professions ( physical therapist and radiologist), and a clearly defined and limited inclusion and assessment period may enhance the reliability of ultrasound examinations. Alternatively, 3D-CT analysis could be utilized to evaluate the position of the humeral head relative to the glenoid cavity in patients experiencing anterior shoulder instability. This evaluation can be effectively combined with the analysis of associated bony lesions of the humeral head or glenoid bone. However, CT scans are costly and not easily used outside hospital settings.

## Conclusion

In patients, a significantly greater anterior translation of the humeral head was observed during active extension of the elbow compared to participants in the asymptomatic control group. The findings of this study further emphasize the difficulties in measuring anterior glenohumeral translation during active motion within a clinically feasible setup. The HHC test demonstrated poor to moderate reliability, with elbow extension demonstrating the highest test-retest and inter-rater reliability. Therefore, this novel test, in its current form, cannot be recommended for use in clinical practice or research.

## Data Availability

All data relevant to the study are included in the article

## Acknowledgment

We thank Prof. Dr. H.E.J. Veger for his critical review of the analysis used in our manuscript.

## Notes

### Competing Interest Statement

I have read the journal's policy and the authors of this manuscript have the following competing interests: M.P.J. van den Bekerom: The OLVG receives fellowship support from Smith and Nephew. All other authors: none

### Clinical Trial

NL-OMON21609

### Author Declarations

The Medical Research Ethics Committees United (MEC-U) Registration number NL69882.100.19.

